# ACE2 levels are altered in comorbidities linked to severe outcome in COVID-19

**DOI:** 10.1101/2020.06.04.20122044

**Authors:** Valur Emilsson, Elias F Gudmundsson, Thor Aspelund, Brynjolfur G Jonsson, Alexander Gudjonsson, Lenore J. Launer, John R Lamb, Valborg Gudmundsdottir, Lori L Jennings, Vilmundur Gudnason

**Author notes:** These authors contributed equally as joint-first authors (Valur Emilsson, Elias F Gudmundsson). Corresponding authors: Valur Emilsson and Vilmundur Gudnason, Icelandic Heart Association, Holtasmari 1, IS-201 Kopavogur, Iceland.

## Abstract

**Aims:** Severity of outcome in COVID-19 is disproportionately higher among the obese, males, smokers, those suffering from hypertension, kidney disease, coronary heart disease (CHD) and/or type 2 diabetes (T2D). We examined if serum levels of ACE2, the cellular entry point for the coronavirus SARS-CoV-2, were altered in these high-risk groups.

**Methods:** Associations of serum ACE2 levels to hypertension, T2D, obesity, CHD, smokers and males in a single center population-based study of 5457 Icelanders from the Age, Gene/Environment Susceptibility Reykjavik Study (AGES-RS) of the elderly (mean age 75±6 years).

**Results:** Smokers, males, and individuals with T2D or obesity have altered serum levels of ACE2 that may influence productive infection of SARS-CoV-2 in these high-risk groups.

**Conclusion:** ACE2 levels are upregulated in some patient groups with comorbidities linked to COVID-19 and as such may have an emerging role as outcome in COVID-19. a circulating biomarker for severity of severity of outcome in COVID-19.

**Key Points:** *Question:* Severity of outcome in COVID-19 is disproportionately higher among the obese, males, smokers, those suffering from hypertension, kidney disease, coronary heart disease (CHD) and/or type 2 diabetes (T2D). Thus, we asked if the coronavirus SARS-CoV-2 receptor ACE2 was altered in the sera from these high-risk groups?

*Findings:* In a single center population-based study of 5457 Icelanders, the Age, Gene/Environment Susceptibility Reykjavik Study (AGES-RS), we find that ACE2 levels are significantly elevated in serum from smokers, obese and diabetic individuals, while reduced in males.

*Meaning:* These results demonstrate that individuals with comorbidities associated with infection of SARS-CoV-2 in these individuals. severe outcome in COVID-19 have altered serum levels of ACE2 that may influence productive

## Introduction

The current coronavirus disease 2019 (COVID-19) pandemic is caused by the severe acute respiratory syndrome coronavirus 2 (SARS-CoV-2)^1^. SARS-CoV-2 and other SARS coronaviruses interact with the receptor ACE2^2^, to enter the host cell. SARS-CoV-2 binds ACE2 with a 10- to 20-fold higher affinity than other SARS coronaviruses do^3^. Furthermore, when the SARS-CoV-2 spike protein S binds ACE2, concomitant shedding of the receptor promotes cellular entry of the virus^4^. COVID-19 is associated with major respiratory failure with highest rates of severe outcome among the elderly, males and/or those with an underlying chronic disease including but not limited to coronary heart disease (CHD), hypertension, kidney disease, type 2 diabetes (T2D) and obesity (kg/m^2^ > 30)^5–7^. This is particularly concerning for the more economically developed countries which have high prevalence of obesity and T2D^8^.

While lower survival in COVID-19 might be attributed to the frailty and vulnerabilities of the high-risk patient groups mentioned above, other underlying factors could play role. For instance, variable levels and/or activity of the membrane bound receptor ACE2^2^ and its soluble counterpart may contribute to this susceptibility. Just how the balance between the levels of soluble ACE2 and its membrane-bound receptor counterpart is maintained, has still to be fully defined. We postulate that productivity of the SARS-CoV-2 infection may vary among individuals depending on their endogenous levels of circulating ACE2. To address this, we examined ACE2 levels in serum from persons with hypertension, obesity, T2D or CHD and other relevant phenotypes, in a population of 5457 individuals aged 65 and above.

## Methods

### Study population

Participants aged 66 through 96 were from the Age, Gene/Environment Susceptibility Reykjavik Study (AGES-RS) cohort^9^, a single-center prospective population-based study aimed to understand aging in the context of gene-environment interactions. The AGES-RS was approved by the NBC in Iceland (approval number VSN-00–063), and by the National Institute on Aging Intramural Institutional Review Board (U.S.) and the Data Protection Authority in Iceland.

### Study design and statistical analysis

The following co-morbidities and characteristics were investigated in multiple regression analysis for an effect on ACE2 levels: age, sex, and body mass index (BMI) (kg/m^2^) categories. Type 2 diabetes was defined as fasting serum glucose > 7.0 mmol/L or self-reported history of diabetes or the use of insulin or oral glucose-lowering drugs. Hypertension was investigated using four different variables; systolic blood pressure > 140 or diastolic blood pressure > 90, use of ARB medications and ACE inhibitors as well as any other hypertension medications. Information on current smoking was obtained from questionnaires. CHD was ascertained using hospital records and/or cause of death information. A CHD event was defined as any occurrence of myocardial infarction, ICD-10 codes: I21-I25, coronary revascularization (either CABG-surgery or PTCA intervention) or death from CHD. Chronic obstructive pulmonary disease (COPD) was defined according to hospital records ICD-10 code: J44, medications and questionnaire. Estimated glomerular filtration rage (eGFR) was calculated from age, sex, and serum creatinine. Means and standard deviations were calculated for continuous characteristics and numbers and percentages for categorical characteristics. The Framingham Risk Score is a multivariable risk function that estimates an individuals 10-year CVD risk^10^.

### Protein measurements

Serum protein levels of ACE2 was measured in 5457 individuals from the AGES-RS using the aptamer-based protein profiling platform^11^. Various metrics related to the performance of the proteomic platform including aptamer specificity, assay variability and reproducibility have already been described^11^. ACE2 was investigated on a log2 scale, where a one-unit increase represents a doubling in levels on the RFU scale. A multiple linear regression with log2-ACE2 levels as the dependent variable was performed with adjustments for the clinical covariates mentioned above. Observed levels of ACE2 were graphically presented by BMI-categories using violin plots. A P-value < 0.05 was considered statistically significant in hypothesis testing.

## Results

Results from multiple linear regression analysis with ACE2 levels (log2) as the outcome are highlighted in **Table 1**. ACE2 serum levels were reduced in serum from males compared to females (β = –0.0385, P < 0.001). In contrast, serum ACE2 levels were increased in overweight (β = 0.0259, P = 0.004) and obese individuals (β = 0.0246, P = 0.036) when compared to lean (BMI < 25) individuals. ACE2 levels measured higher in severely obese (β = 0.0331, P = 0.087), but did not reach significance most likely because few individuals are severely obese in this cohort of old people. The violin plots in **Figure 1** demonstrate elevated levels (log2 scale) of ACE2 in serum in response to increasing adiposity presented by different BMI categories. Both individuals with impaired fasting glucose levels (β = 0.0295, P < 0.001) and those with established T2D (β = 0.0377, P = 0.003) showed higher ACE2 levels compared to those with no T2D or with normal glucose levels (**Table 1**). Serum levels were significantly increased in current smokers (β = 0.0299, P = 0.018), but were not associated with CHD, COPD or estimated glomerular filtration rate (eGFR) (**Table 1**). Given many of the clinical traits and covariates listed in **Table 1** are components of the Framingham risk score (FRS), we examined the association of the FRS with ACE2 levels. Here, ACE2 serum levels were positively associated with the FRS (β = 0.0726, P = 0.008).

**Figure 1.**
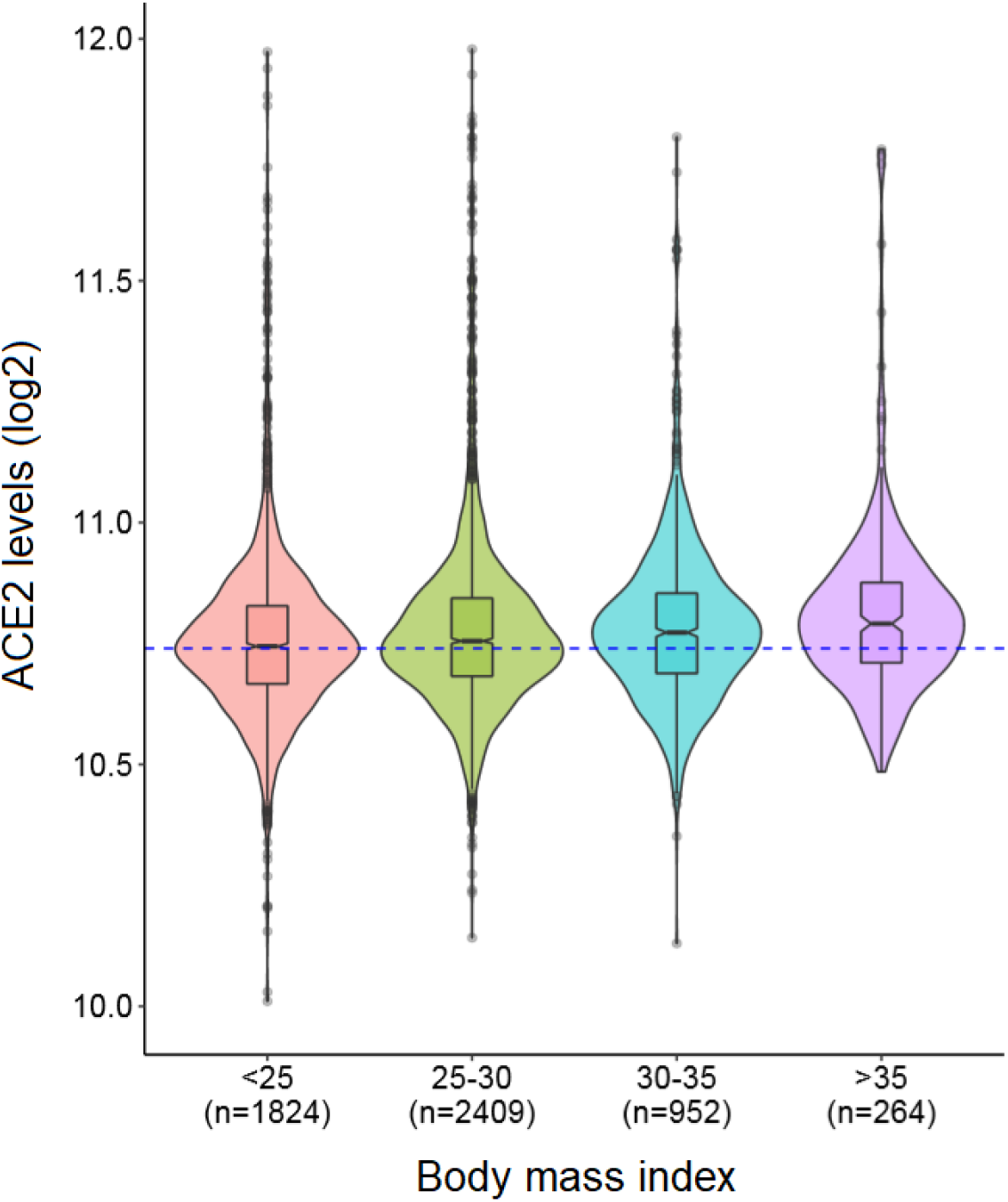
The y-axis represents log2-transformed distribution of ACE2 levels in the serum while the x-axis shows different categories of BMI (kg/m^2^), also expressed in Table 1.

**Table 1.**
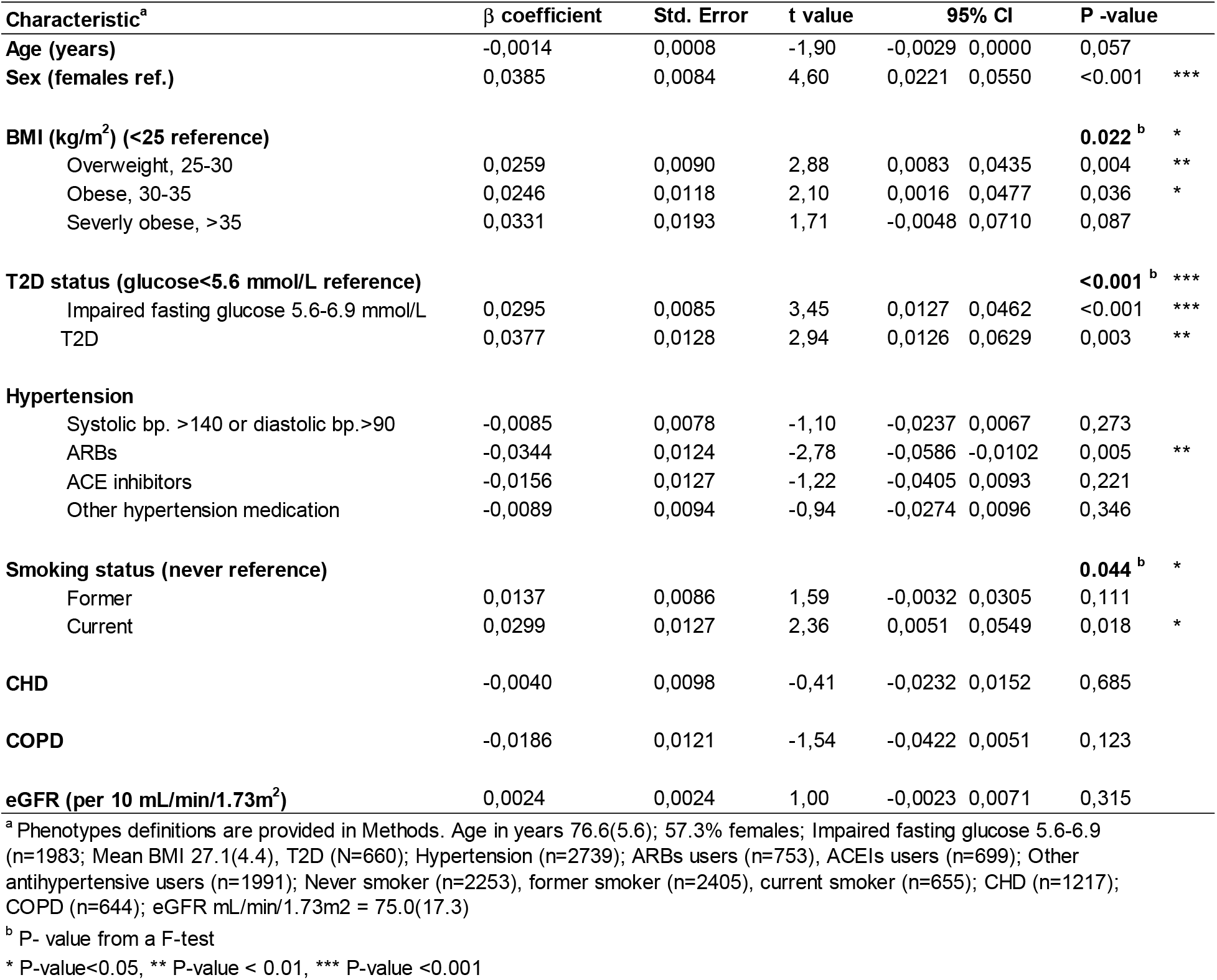
Results from a multiple linear regression analysis with ACE2 levels (log2) as the dependent variable.

| Characteristic<sup>a</sup> | $\beta$ coefficient | Std. Error | t value | 95% CI | | P -value |
| --- | --- | --- | --- | --- | --- | --- |
| **Age (years)** | -0,0014 | 0,0008 | -1,90 | -0,0029 | 0,0000 | 0,057 |
| **Sex (females ref.)** | 0,0385 | 0,0084 | 4,60 | 0,0221 | 0,0550 | <0.001 *** |
| **BMI (kg/m<sup>2</sup>) (<25 reference)** |  |  |  |  |  | **0.022<sup>b</sup> \*** |
| Overweight, 25-30 | 0,0259 | 0,0090 | 2,88 | 0,0083 | 0,0435 | 0,004 ** |
| Obese, 30-35 | 0,0246 | 0,0118 | 2,10 | 0,0016 | 0,0477 | 0,036 * |
| Severely obese, >35 | 0,0331 | 0,0193 | 1,71 | -0,0048 | 0,0710 | 0,087 |
| **T2D status (glucose<5.6 mmol/L reference)** |  |  |  |  |  | **<0.001<sup>b</sup> \*\*\*** |
| Impaired fasting glucose 5.6-6.9 mmol/L | 0,0295 | 0,0085 | 3,45 | 0,0127 | 0,0462 | <0.001 *** |
| T2D | 0,0377 | 0,0128 | 2,94 | 0,0126 | 0,0629 | 0,003 ** |
| **Hypertension** |  |  |  |  |  |  |
| Systolic bp. >140 or diastolic bp.>90 | -0,0085 | 0,0078 | -1,10 | -0,0237 | 0,0067 | 0,273 |
| ARBs | -0,0344 | 0,0124 | -2,78 | -0,0586 | -0,0102 | 0,005 ** |
| ACE inhibitors | -0,0156 | 0,0127 | -1,22 | -0,0405 | 0,0093 | 0,221 |
| Other hypertension medication | -0,0089 | 0,0094 | -0,94 | -0,0274 | 0,0096 | 0,346 |
| **Smoking status (never reference)** |  |  |  |  |  | **0.044<sup>b</sup> \*** |
| Former | 0,0137 | 0,0086 | 1,59 | -0,0032 | 0,0305 | 0,111 |
| Current | 0,0299 | 0,0127 | 2,36 | 0,0051 | 0,0549 | 0,018 * |
| **CHD** | -0,0040 | 0,0098 | -0,41 | -0,0232 | 0,0152 | 0,685 |
| **COPD** | -0,0186 | 0,0121 | -1,54 | -0,0422 | 0,0051 | 0,123 |
| **eGFR (per 10 mL/min/1.73m<sup>2</sup>)** | 0,0024 | 0,0024 | 1,00 | -0,0023 | 0,0071 | 0,315 |

<sup>a</sup> Phenotypes definitions are provided in Methods. Age in years 76.6(5.6); 57.3% females; Impaired fasting glucose 5.6-6.9 (n=1983; Mean BMI 27.1(4.4), T2D (N=660); Hypertension (n=2739); ARBs users (n=753), ACEIs users (n=699); Other antihypertensive users (n=1991); Never smoker (n=2253), former smoker (n=2405), current smoker (n=655); CHD (n=1217); COPD (n=644); eGFR mL/min/1.73m<sup>2</sup> = 75.0(17.3)
<sup>b</sup> P- value from a F-test
\* P-value<0.05, \*\* P-value < 0.01, \*\*\* P-value <0.001

## Discussion

The severity of outcome in COVID-19 is greatly influenced by various comorbidities, unhealthy lifestyle and being a male^5–7^. We observed an increase in circulating levels of ACE2 in some of these groups including smokers, obese and diabetics and found that ACE2 levels were positively correlated with the FRS. In contrast, ACE2 levels were decreased in serum from males compared to females, thus showing an opposite effect from that of the comorbidities associated with COVID-19. Intriguingly, studies have shown that estrogen levels may induce ACE2 activity and levels in females^12^. However, these results do not agree with a recent study showing higher plasma levels of ACE2 in male patients suffering from heart failure^13^. Our observation that ACE2 levels were elevated in smokers agrees well with a study showing upregulation of ACE2 mRNA in the respiratory tract of smokers as well as rodents exposed to cigarette smoke^14^.

ACE2 is a membrane-bound ectoenzyme that is released into the circulation *via* ectodomain shedding^15^. Variable levels of ACE2 in serum is likely a result of genetic factors, differential gene expression and/or ectodomain shedding influenced by disease or administration of drugs *via* negative or positive feedback loops of the renin-angiotensin system^16^. As proteins in circulation emanate from virtually every tissue of the body^11^, we cannot easily determine how much each tissue contributes to ACE2 release into blood. For instance, high serum ACE2 levels may not mirror higher activity or abundances of its membrane bound counterpart in pulmonary tissues. However, we show that serum levels of ACE2 are altered in some frequent comorbidities and other related phenotypes associated with severity of outcome in COVID-19.

## Limitations

The findings showing higher ACE2 levels in the serum of smokers, diabetic and obese individuals warrant further validation in independent study populations. This work does not show that altered ACE2 serum levels in these patient groups causes worse clinical outcomes in COVID-19, though that is a possibility. The results presented may be limited to that of serum and may not reflect the effect of these comorbidities on ACE2 levels in pulmonary tissues, but lung injury is the key component linked to outcome severity in COVID-19. Furthermore, the measured circulating levels of ACE2 may not reflect its cell surface receptor levels in solid tissues. Finally, all participants of the AGES-RS are white (Caucasians) which may limit the transferability and generalizability of the results as ACE2 levels differ across races and ethnicities^17^.

## Conclusions

Individuals that are males, smokers, manifest with diabetes or obesity are at high risk of developing severe outcome from COVID-19. In case of the smokers, obese and diabetics, the increased risk could partly be explained by the vulnerability of these patient groups. We find that serum levels of ACE2 are altered in all these groups. However, while serum ACE2 levels were increased in smokers, diabetic and obese individuals, males had reduced ACE2 serum levels. These results encourage further efforts in defining the relationship between soluble ACE2 levels and the membrane bound ACE2 form in solid tissues.

## Data Availability

The custom-design Novartis SOMAscan is available through a collaboration agreement with the Novartis Institutes for BioMedical Research. Data from the AGES Reykjavik study are available through collaboration under a data usage agreement with the IHA

## Acknowledgment

The present study was supported by Novartis Institute for Biomedical Research (NIBR), Icelandic Heart Association, and in part by the intramural research program at the National Institute of Aging (N01-AG-12100 and HHSN271201200022C). We thank all AGES-Reykjavik study participants, and the staff of the IHA for their contribution to the AGES-Reykjavik study. The custom-design Novartis SOMAscan is available through a collaboration agreement with the Novartis Institutes for BioMedical Research. Data from the AGES Reykjavik study are available through collaboration under a data usage agreement with the IHA. J.R.L. and L.L.J. were and are, respectively, employees and stockholders of Novartis. All other authors declare they have no competing interests.

## References

1. Lu H, Stratton CW, Tang YW. Outbreak of pneumonia of unknown etiology in Wuhan, China: The mystery and the miracle. J Med Virol. 2020;92(4):401–402.

2. Wan Y, Shang J, Graham R, Baric RS, Li F. Receptor Recognition by the Novel Coronavirus from Wuhan: an Analysis Based on Decade-Long Structural Studies of SARS Coronavirus. J Virol. 2020;94(7).

3. Wrapp D, Wang N, Corbett KS, et al. Cryo-EM structure of the 2019-nCoV spike in the prefusion conformation. Science. 2020;367(6483):1260–1263.

4. Ziegler CGK, Allon SJ, Nyquist SK, et al. SARS-CoV-2 Receptor ACE2 Is an Interferon-Stimulated Gene in Human Airway Epithelial Cells and Is Detected in Specific Cell Subsets across Tissues. Cell. 2020.

5. Zhou F, Yu T, Du R, et al. Clinical course and risk factors for mortality of adult inpatients with COVID-19 in Wuhan, China: a retrospective cohort study. Lancet. 2020;395(10229):1054–1062.

6. Richardson S, Hirsch JS, Narasimhan M, et al. Presenting Characteristics, Comorbidities, and Outcomes Among 5700 Patients Hospitalized With COVID-19 in the New York City Area. Jama. 2020.

7. Preliminary Estimates of the Prevalence of Selected Underlying Health Conditions Among Patients with Coronavirus Disease 2019 – United States, February 12-March 28, 2020. MMWR Morb Mortal Wkly Rep. 2020;69(13):382–386.

8. Hossain P, Kawar B, El Nahas M. Obesity and diabetes in the developing world--a growing challenge. N Engl J Med. 2007;356(3):213–215.

9. Harris TB, Launer LJ, Eiriksdottir G, et al. Age, Gene/Environment Susceptibility-Reykjavik Study: multidisciplinary applied phenomics. Am J Epidemiol. 2007;165(9):1076–1087.

10. D’Agostino RB, Vasan RS, Pencina MJ, et al. General Cardiovascular Risk Profile for Use in Primary Care. Circulation. 2008;117(6):743–753.

11. Emilsson V, Ilkov M, Lamb JR, et al. Co-regulatory networks of human serum proteins link genetics to disease. Science. 2018;361(6404):769–773.

12. Zhang Q, Cong M, Wang N, et al. Association of angiotensin-converting enzyme 2 gene polymorphism and enzymatic activity with essential hypertension in different gender: A case-control study. Medicine (Baltimore). 2018;97(42):e12917.

13. Sama IE, Ravera A, Santema BT, et al. Circulating plasma concentrations of angiotensin-converting enzyme 2 in men and women with heart failure and effects of reninangiotensin-aldosterone inhibitors. Eur Heart J. 2020;41(19):1810–1817.

14. Smith JC, Sausville EL, Girish V, et al. Cigarette smoke exposure and inflammatory signaling increase the expression of the SARS-CoV-2 receptor ACE2 in the respiratory tract. Dev Cell. 2020.

15. Lambert DW, Yarski M, Warner FJ, et al. Tumor necrosis factor-alpha convertase (ADAM17) mediates regulated ectodomain shedding of the severe-acute respiratory syndrome-coronavirus (SARS-CoV) receptor, angiotensin-converting enzyme-2 (ACE2). J Biol Chem. 2005;280(34):30113–30119.

16. Patel SK, Velkoska E, Freeman M, Wai B, Lancefield TF, Burrell LM. From gene to protein-experimental and clinical studies of ACE2 in blood pressure control and arterial hypertension. Front Physiol. 2014;5:227.

17. Ciaglia E, Vecchione C, Puca AA. COVID-19 Infection and Circulating ACE2 Levels: Protective Role in Women and Children. Front Pediatr. 2020;8:206.

